# COBT: A gene-based rare variant burden test for case-only study designs using aggregated genotypes from public reference cohorts

**DOI:** 10.1101/2025.07.16.25331553

**Authors:** Antoine Favier, Stefania Chounta, Alejandro Garcia, Fabienne Jabot-Hanin, Xiaoyi Chen, Nicolas Garcelon, Anita Burgun, Manuel Higueras, Agathe Guilloux, Alexandre Benmerah, Yoann Martin, Katy Billot, Jean-Michel Rozet, Isabelle Perrault, Valérie Cormier-Daire, Céline Huber, Mohamad Zaidan, Tania Attie-Bitach, Sophie Saunier, Antonio Rausell

## Abstract

More than 4000 rare genetic diseases affect 1 in 16 people, yet ∼50% of patients remain undiagnosed after genetic testing. Identifying genotype-phenotype associations is challenged by small cohorts and high clinical and genetic heterogeneity. Rare variant burden tests increase statistical power in case-control studies, but are limited in rare disease research due to the lack of matched controls in retrospective studies. Recently proposed case-only aggregation tests assess the number of individuals with damaging variants under dominant or recessive models, but fail to capture additive effects, hypomorphic variants, or heterogeneous inheritance modes. Here, we present the Case-Only Burden Test (COBT), a gene-based burden test for case-only designs accounting for multiple variants per individual and their putative additive effects. COBT uses a Poisson model to test for excess variants in a gene compared to expectations from general population mutation rates. We validated the model’s assumptions and goodness-of-fit on a control cohort from the 1000 Genomes Project, where COBT showed low false-positive rates and outperformed alternative case-only tests. Applied to 478 ciliopathy patients, COBT re-identified known causal genes and highlighted novel candidate variants in unsolved cases. COBT enables gene discovery in case-only rare disease cohorts and is available at https://github.com/RausellLab/COBT.

## Background

Current diagnostic rates of Mendelian rare disease patients are heterogeneous across disease types (1). In the case of ciliopathies, a group of disorders resulting from abnormal formation or function of cilia, between 25% and 40% of patients remain without a pathophysiological label after genetic testing (2, 3). Molecular diagnosis is challenged by a large degree of both genetic and phenotypic heterogeneity across individuals (4–6). From a clinical point of view, both allele and loci heterogeneity have been described, where different mutations within the same or different genes may cause the same clinical outcomes. From a genetic standpoint, the same or different variants in a given gene may lead to heterogeneous consequences across patients in terms of variable severity — where the same phenotype may manifest at diverse intensities — but also in the form of variable expressivity, where differing phenotypes may be observed. Heterogeneous effect sizes of disease-associated variants as well as the interplay of additive effects, including modifier variants with susceptibility or protective effects (7), have been proposed to explain the observed clinical heterogeneity across patients, in addition to environmental factors and the genetic background such ethnicity and sex (8). From a statistical genetics’ perspective, such genetic and clinical heterogeneity may compromise the statistical power to identify significant genotype-phenotype associations in rare disease cohorts, ultimately leading to important rates of genetically uncharacterized patients for some pathologies (9).

In certain cases, however, such limitation may be uncovered through burden tests of genetic variants in case-control study designs. Here, the number of variants carried by an individual in a specific genomic region is evaluated in a weighted aggregated manner against the presence / absence of a phenotype across individuals. A diversity of burden tests has been developed using different statistical hypothesis, genomic aggregation elements (e.g. coding exons, entire genes or set of genes), qualifying variant types (*i*.*e*., the characteristics of genetic variants considered by the test depending on their predicted molecular consequences or inheritance mode) as well as weighting schemes, where the contribution of a variant to the aggregated statistic is rated according to its pathogenicity potential through proxies such allele frequency or bioinformatics scores (10). While having proved a successful strategy for identifying novel gene candidates, the use of burden tests in rare disease studies is in practice often limited by the lack of matched control samples in retrospective cohort study designs.

To overcome such limitation, rare variant aggregation tests such as TRAPD (11) and CoCoRV (12) have been proposed for case-only study designs. These methods evaluate whether there is an excess on the number of patients bearing at least one qualifying variant in a given gene or genomic region, as compared to the expectations that would be obtained if the same number of individuals were randomly sampled from the general population. To that aim, large-scale population sequencing data from publicly available databases such gnomAD have been used as a reference (13). The observed and expected number of carrier individuals are then compared through statistical tests such as Chi-squared, Fisher exact and Cochran–Mantel–Haenszel (CMH)-exact tests. By doing so, TRAPD and CoCoRV evaluate the number of carrier individuals rather than the variant burden within individuals and cohorts, which poses major limitations. Thus, they rely on a sharp definition of the variant type that would make an individual being considered as carrying pathogenic variants or not, which may lead to neglecting hypomorphic and modifier variants (7). Second, those methods cannot easily accommodate genetic architectures presenting with heterogeneous Mendelian inheritance patterns (*i*.*e*., >7% of rare genetic diseases) (14), including incomplete dominance, co-dominance and compound heterozygosity. Finally, those tests are insensitive to the presence of multiple variants within an individual and their eventual additive effects, which is a major characteristic of burden tests in case-control designs (8).

To fill this gap, in this study we present the COBT method (Case-Only Burden Test), the first rare variant burden test for case-only study designs able to account both for the presence of multiple variants within individuals and their putative additive effects. COBT evaluates the null hypothesis stating that the total number of qualifying variants in a given gene observed across the patients in a given cohort is not statistically different from the number that might be expected based on the gene mutation rates observed in the general population. Such hypothesis is tested through a Poisson test parametrized on the gene mutation rates observed in the general population, using gnomAD database as a reference. We first characterized COBT’s behaviour on a target cohort of the general population, constituted by non-Finnish European individuals from the 1000 Genomes Project, where no rejections of the null hypothesis were *a priori* expected. Such cohort served as a negative control upon which we performed the evaluation of the statistical assumptions and goodness-of-fit of the test, the diagnosis of its p-value distributions and inflation rates, as well as an in-depth analysis of the test sensitivity to false positive detection. We then applied COBT to a case-only cohort of 478 patients suffering from diverse ciliopathy types in order to evaluate its ability to recapitulate known causal genes previously identified in the cohort, as well as to identify novel candidate causal variants that may be proposed as primary or secondary hits for the genetically unsolved cases.

## Methods

### Sequencing data

Two *target* cohorts were considered. First, whole-genome sequencing data from non-Finnish European individuals from the 1000 Genomes project Phase 3 (15), based on genome reference GRCh37/hg19 version, were downloaded in Variant Calling Format from http://ftp.1000genomes.ebi.ac.uk/vol1/ftp/release/20130502/. Second, ciliary exome-targeted sequencing, referred in the text as *ciliome sequencing*, was conducted in an in-house cohort of 478 patients suffering from ciliopathies, including nephronophthisis-related renal ciliopathies, retinal ciliopathies, skeletal ciliopathies, and embryonic and foetal ciliopathies, under Ethical approval committee *Comité Éthique et Scientifique pour les Recherches, les Études et les Évaluations dans le domaine de la Santé (CESREES)*, approved on September 3^rd^, 2020, number #2201437. Patients included 224 females and 243 males (11 unknown), from diverse ethnicity origins. Genomic DNA was isolated from blood lymphocytes and subjected to exome capture using a custom SureSelect capture kit (Agilent Technologies) targeting 4.5 Mb of 20,168 exons (1,221 ciliary candidate genes) (16, 17). Sequencing performed on SOLiD5500XL (Life Technologies) and HiSeq2500 (Illumina) was done on pools of barcoded ciliome libraries. Paired-end reads were generated (75+35 for SOLiD, 100+100 for HiSeq) and sequences were aligned to the reference human genome hg19 with Illumina’s processing software ELAND (CASAVA 1.8.2), the Burrows–Wheeler Aligner (Illumina) or mapread (SoliD). Downstream processing was carried out with the Genome Analysis Toolkit (GATK), SAMtools, and Picard Tools, following documented best practices (https://gatk.broadinstitute.org/hc/en-us/sections/360007226651-Best-Practices-Workflows). Variants were jointly called with GATK4 Haplotypecaller. As the *reference* cohort representing the general population, exome sequencing data from the Genome Aggregation Database (gnomAD) was used throughout the study (Version 2.1, hail tables available from https://gnomad.broadinstitute.org/downloads). Only non-Finnish europeans samples from gnomAD (n=56,885) were considered for the evaluation of the 1000 Genomes data, whereas all samples (n=125,748) were used for the ciliome analysis.

### Genomic regions and sample filtering

Analyses were restricted throughout the study to human protein-coding genes mapping autosomal chromosomes as obtained from BioMart Ensembl release 108, GRCh37.p13 (18). Protein-coding genes were defined as those containing an open reading frame. In addition, previously-described signal-artifact blacklisted regions of the human genome (19) (as provided at https://github.com/BoyleLab/Blacklist/raw/master/lists/hg19-blacklist.v2.bed.gz) were filtered out. Blacklisted regions were assessed with bioinformatics tools based on mappability: anomalous, unstructured, high signal/read counts in independent cell-lines from ENCODE and then curated by hand. Furthermore, a gene exclusion list was considered, including 2,157 genes with highly polymorphic regions and characteristics of assembly misalignments identified in exome data from 118 individuals from 29 families (20), obtained from https://onlinelibrary.wiley.com/action/downloadSupplement?doi=10.1002%2Fhumu.22033&file=Table_S7_gene_exclusion_list_final.txt. For genomic coordinates considerations, only the principal isoform for each gene were considered, as provided by APPRIS database (21) based on GRCh37/hg19 and Gencode v19 (https://appris.bioinfo.cnio.es/#/downloads). In addition, as suggested for rare-variant collapsing analyses (10), genomic regions were further restricted to those mapping so-called ‘trustworthy regions’, defined as those covered at 10X or more in at least 90% of the samples in both the reference (*i*.*e*., gnomAD) and the target cohort (either the 1000 Genomes or the ciliome cohort described above). In the case of the 1000 Genomes data, analyses were further restricted to regions passing “strict mask” filters, as provided at http://ftp.1000genomes.ebi.ac.uk/vol1/ftp/release/20130502/supporting/accessible_genome_masks/StrictMask/, and defined as having a total coverage within 50% of the average, having no more than 0.1% of reads with mapping quality of zero, and with an average mapping quality for the position equal or greater than 56. Genes covered in less than 25% of the APPRIS Principal transcript length after intersection among ‘trustworthy regions’ were filtered out. Samples with genotype quality < 20, depth < 8 or < 95% call-rate (percentage of variants for which no genotype could confidently be called) were discarded. A schematic representation of the above mentioned genomic and sample filtering is provided in **Supplementary figure 1**.

### Genetic variant annotation and filtering

Single nucleotide variants (SNVs) and indels mapping the retained genomic regions described above were considered. Throughout the study, custom variant filtering was performed with hail library 0.2.57 (22). Variant annotation was performed using VEP (23) v100.4. Only variants with Allele Frequency (AF) < 1% in the target cohort and < 0.1% in the reference cohort were retained. Only variants annotated as synonymous, missense or Protein Truncating Variant (PTV) on the APPRIS principal isoform for each gene were considered (21). PTVs were defined as those annotated as stop-gained, splice acceptor, splice donor and frameshift variants. When a variant mapped 2 overlapping genes, the gene/transcript with the most damaging consequence was retained, with the following preference: PTV > missense variant > synonymous variant. Protein altering variants were defined as those annotated either as missense or PTV. A schematic representation of the above mentioned genetic variant annotation and filtering is provided in **Supplementary figure 1**.

### Goodness-of-fit Poisson tests

Let X={*x*_1_, *x*_2_, …, *x*_*n*_} represent a sample of *n* independent counts, and 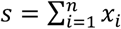 its sample sum. Several statistical tests allow to determine whether they derive from a Poisson distribution. First, the *D*-test is an exact test evaluating the dispersion of the data (24). The test is a two-tailed evaluation of the cumulative exact probability of the sum of squares of the supposed Poisson realizations and is derived from Fischer’ exact D-test (25):

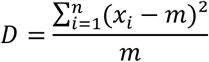

Second, the *L*-test (26) evaluates the homogeneity of the data, *i*.*e*., whether a set of Poisson realizations have the same parameter. The *L*-test is a one-tailed test and is based on the likelihood ratio test. The L-statistic is assessed as follows:

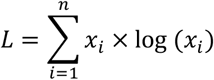

P-values for testing dispersion (and homogeneity) are assessed by computing the cumulative probability of the realizations of greater or equal *D* (or *L* for the homogeneity) (27).

Finally, the CR-test (24) compares the frequency of zeros observed in the sample with the distribution of zeros expected from the Poisson assumptions, *N*_0_. Considering *n*_0_, the sample number of observed zeros, one can compute the right-tailed CR-test p-value for the null-hypothesis stating that the data is Poisson distributed against the alternative hypothesis that data is zero-inflated:

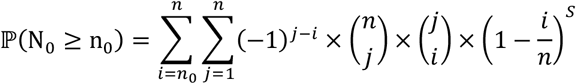

For each test, significance was assessed after Bonferroni correction. The code to run the previously described tests was adapted from a shiny app available online (https://manu2h.shinyapps.io/gof_poisson/) and described in Fernández-Fontelo *et al*. (24).

### Inflation factor estimation

Inflation factor estimation was performed using two alternative expectations about the p-values distribution under the null hypothesis H_o_. First, assuming a uniform distribution of p-values, and second, empirically sampling the null Poisson distribution under the null hypothesis. Here, following Higueras *et al*. (24, 27), for each evaluated gene, we randomly sampled the number of variants from a Poisson distribution with parameter λ, and calculated their associated Poisson p-values. We did so N independent times. For each simulation, we sorted the P-values under null across all genes and stored their rank. Finally, the expected sorted P value at rank k was assessed as the average of the p-values having ranked in position k across each of the N simulations. To estimate the inflation factor, we took the lower 95% quantile of points in the QQ plot and regressed the sorted log10-scaled observed P values to the log10-scaled expected sorted P values. The slope of the regression against the raw p-values was defined as the inflation factor, denoted by 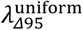 or 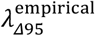, depending on whether the uniform distribution or the sampled-null distribution was used, respectively. When inflation factors were assessed upon genomic corrected p-values, the analogous figures where denoted by 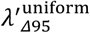 or 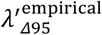.

### Genomic correction of p-values and multiple testing correction

To minimize the effects of confounding factors and especially invisible substructure of the data, genomic control was applied to minimize false positive associations. Following Devlin *et al*. (28), a χ^2^-test statistic with 1 degree of freedom is computed at each locus and λ is estimated as the median of the χ^2^-test divided by 0.456 (*i*.*e*., χ^2^-test with 1 degree of freedom for a p-value of 0.5). New χ^2^-test statistics are divided by λ and p-values for a χ^2^-test statistic with 1 degree of freedom are computed at each locus. P-values are corrected for multiple testing by Bonferroni correction.

### Reference databases of pathogenic variants

The relevance of the most damaging variant of patients mutated in the case-only enriched genes was assessed using two variant pathogenicity databases: ClinVar (29) or the Human Gene Mutation Database (HGMD) (30). CADD (31) was also used to discriminate variants. The most damaging variant of patients were chosen following these criteria: (i) Presence of a pathogenic variant registered in ClinVar or a damaging variant in the Human Gene Mutation Database. (ii) Presence of a probably pathogenic variant registered in ClinVar (pathogenic/likely pathogenic, likely pathogenic) or a probably damaging variant in HGMD (probably damaging, disease-associated polymorphism). (iii) If no record existed in the previous databases, the variant with the highest CADD PHRED score was kept. An arbitrary CADD threshold of 15 was used to define pathogenicity, in accordance with the median value of all possible canonical splice sites and non-synonymous variants of CADD v1.0 (32).

## Results

### Formulation of the case-only gene-based rare variants burden test (COBT)

The distribution of the total number *n*_*j*_ ∈ ℕ of qualifying rare variants identified in an individual *j* across *i* qualifying genes (*i* = 1 … *I*) within a genome may be modelled as a multinomial distribution as follows: let *n*_*j*_ be the number of independent trials (*i*.*e*., genetic variants) each leading to a success for exactly one of mutually exclusive *I* categories (*i*.*e*., genes), with each category having a given normalized fixed-success probability *p*_*i*_, where 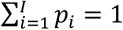. Let 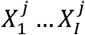 be *I* random variables representing the non-negative integer number of such successes for each category *i*. Thus, the probability of observing any particular combination of numbers of successes 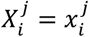 at individual *j* for the various genes *i* is given by the multinomial distribution described by:

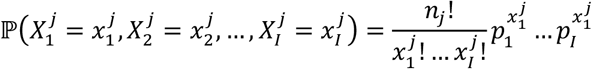

where 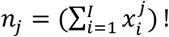

Focusing on a specific gene *i*, such distribution may be simply described as a binomial distribution (**Figure 1**), with parameters *n*_*j*_ and *p*_*i*_, *i*.*e*., 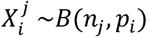, where:

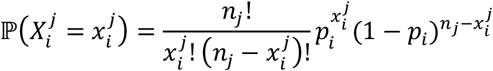

**Figure 1:**
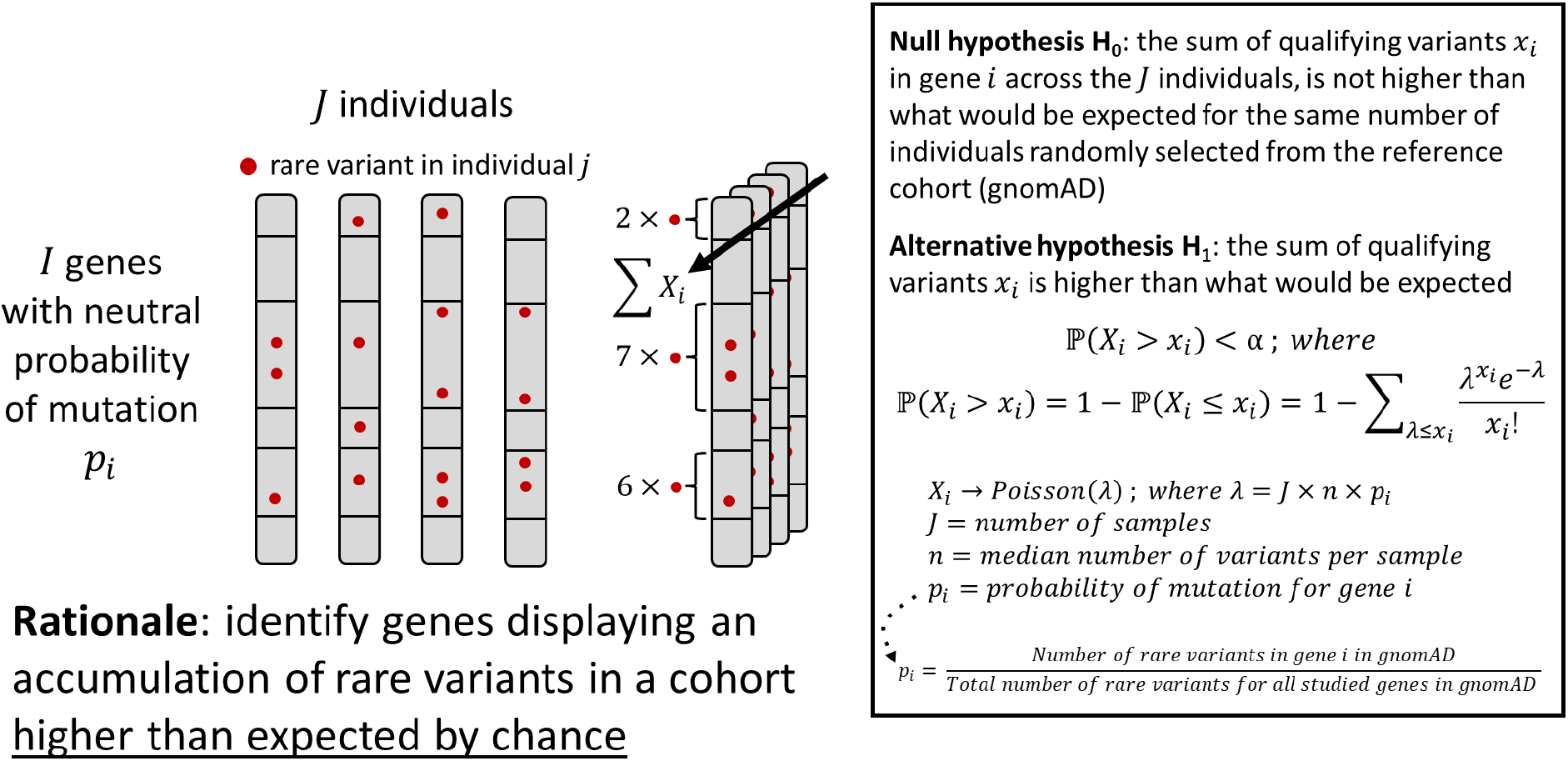
Schematic diagram of the COBT approach. Schematic representation of the concept and statistical model behind COBT.

If multiple individuals are considered, the sum of variants for a specific gene *i* across *J* individuals, represented as *X*_*i*_, may be modelled as a random variable resulting from the sum of the realizations of *J* independent binomial distributions *B*(*n*_*j*_, *p*_*i*_), which behave in turn as a binomial distribution:

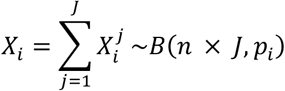

where *n* represents the median of *n*_*j*_ across *J* (**Figure 1**).

When the number *J* of individuals is high, such binomial distribution may be approximated by a Poisson distribution:

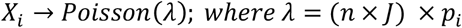

The availability of large-scale genome sequencing projects of the general population provides the opportunity to parametrize the fixed success normalized probabilities *p*_*i*_ from the observed distribution of qualifying variants across qualifying genes. Here, the total number of rare synonymous and missense variants observed in a given gene across individuals from the general population were shown to highly correlate with gene length (33). Thus, probabilities *p*_*i*_ can be defined as:

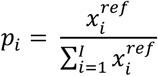

where 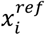 represents the number of unique qualifying variants observed in gene i in a reference cohort. In the present work, gnomAD was used as a *reference* cohort throughout the study (**Methods**).

The previous formalisms allow to evaluate the null hypothesis H_0_ stating that the sum of qualifying variants on a specific gene *i* observed across the *J* individuals from a *target* cohort, *x*_*i*_, is not higher than what would be expected if the same number of individuals would be randomly selected from the *reference* cohort. Thus, by establishing a type I error *α*, the H_0_ rejection is expressed as follows:

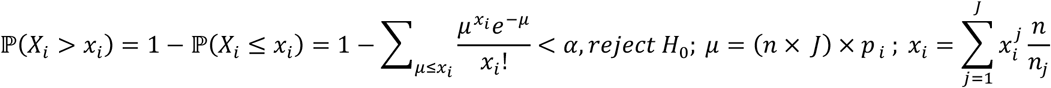

In the previous formula, qualifying variant counts 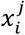 are normalized to account for heterogeneous *n*_*j*_ across individuals, and *x*_*i*_ rounded to the closest integer number.

An additive genetic model was considered throughout this work by which 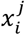 represented the total burden of mutated genomic positions in gene *i* at individual *j*. Thus, heterozygous variants within an individual contributed with one count, while homozygous variants contributed with two counts. However, as done in other rare-variant burden tests in case-control study designs (8), the case-only burden test could be readily adapted to a dominant model without loss of generality, simply by adapting the assessment of 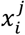 as the number of qualifying variants for which individual *j* carries at least one minor allele.

### Goodness-of-fit of the Poisson distribution for rare variants

We first characterized the behaviour of COBT on a *target* cohort constituted of 404 non-Finnish European individuals from the 1000 Genomes Project (Methods). As for the *reference* cohort, which is based on 56,885 non-Finish Europeans from gnomAD, this *target* cohort represents a random sampling from the general population. As such, it served as a negative control for the purpose of this study as, in principle, no rejections of the null hypothesis H_0_ stated above would be expected. After stringent genomic region filtering (removing error-prone and association tests’ recurrent false positives genomic regions, see Methods), a total of 12,041 qualifying protein-coding genes were retained for downstream analyses, with a total of 9,266 and 10,620 genes presenting at least one qualifying synonymous and missense variant, respectively, in the cohort. We analysed separately the distribution of rare missense and synonymous variants across the *trustworthy* regions of such genes (**Methods**), with a median number per individual of n=80 and n=135 qualifying variants.

First, we inspected whether the distribution of qualifying variants across individuals and qualifying genes approximated the assumption of a Poisson distribution, *i*.*e*., having a variance to mean ratio close to 1. Here, synonymous and missense variants presented per-gene median values of 0.998 and 0.995, with 10^th^ and 90^th^ percentiles of [0.988-1.000] and [0.980-1.000], respectively. The goodness-of-fit of the Poisson-distribution to the observed data was further statistically evaluated in terms of its dispersion, homogeneity and frequency of zeros through the D-test, L-test and CR-test, respectively (Methods). In the case of synonymous variants, a residual 0.4%, 0.5% and 0.2% of the evaluated genes rejected the D, L and CR-test respectively. Similar figures were obtained in the case of missense variants, with 0.1%, 0.2% and 0.1% rejections (**Table 1**). From the previous results, it can be concluded that the per-gene distribution of rare variants across individuals may be modelled through a Poisson distribution for most of the genes, thus supporting the assumptions of the COBT approach.

**Table 1:** Evaluation of the Goodness-of-fit of the Poisson distribution for rare variants. The table shows the number of genes rejecting the D, L and CR tests evaluating the goodness-of-fit of the Poisson distribution in terms of dispersion, homogeneity and zero-inflation, respectively, of the per-gene distribution of qualifying synonymous and missense variants across 404 non-Finnish Europeans from the 1000 Genomes Project.

|  | Synonymous variants | Missense variants |
| --- | --- | --- |
| # genes evaluated (D-test & L-test) | 6,646 | 8,851 |
| # genes evaluated (CR-test) | 9,266 | 10,620 |
| # genes rejecting the D-test | 24 | 12 |
| # genes rejecting the L-test | 30 | 18 |
| # genes rejecting the CR-test | 23 | 16 |
| # genes rejecting the 3 tests | 17 | 9 |

### Diagnosis of p-values distribution and inflation estimation

We then evaluated the distribution of p-values obtained when applying COBT on the target cohort of 404 non-Finnish European individuals from the 1000 Genomes Project. As previously pointed out, the null hypothesis H_0_ of COBT is expected to be true for all genes in this target cohort. Such expectation permits to assess whether the test’s p-value distribution is indeed uniform or, alternatively, to diagnose systematic p-value distribution shifts (34). The inspection of p-value histograms showed a non-uniform distribution of p-values both for synonymous and for missense variants, regardless of whether a genomic correction of p-values was or was not applied (**Supplementary figure 2**).

The associated Quantile-Quantile (QQ)-plots further reflected a systematic inflation of p-values as compared to the expectations from a uniform distribution, with 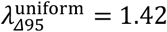 for synonymous variants and 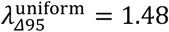 for missense variants (**Supplementary figure 3**). These results are compatible with recent works pointing out that the assumption by which the null distribution of p-values follows a uniform distribution is not well suited for statistical tests based on discrete count data (12, 35, 36). Following (37) we thus sampled the true null distribution of the discrete test statistics in order to obtain a more realistic estimation of the expected distribution of p-values (Methods). Doing so allowed a more accurate inflation factor estimation method, yet, reflecting a systematic inflation(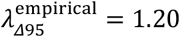 and 1.26 for synonymous and missense variants, respectively) and a significant deviation of p-values from the regression line (**Supplementary figure 3**). However, applying a genomic control of p-values (28, 38) (Methods) corrected for global systematic inflation as well as for deviation of observed p-values from the fitted regression lines (**Figure 2**), and this assuming a uniform (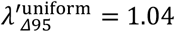 and 1.15, for synonymous and missense variants, respectively) as well as an empirical sampled-null distribution (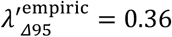 and 0.40, for synonymous and missense variants, respectively).

**Figure 2:**
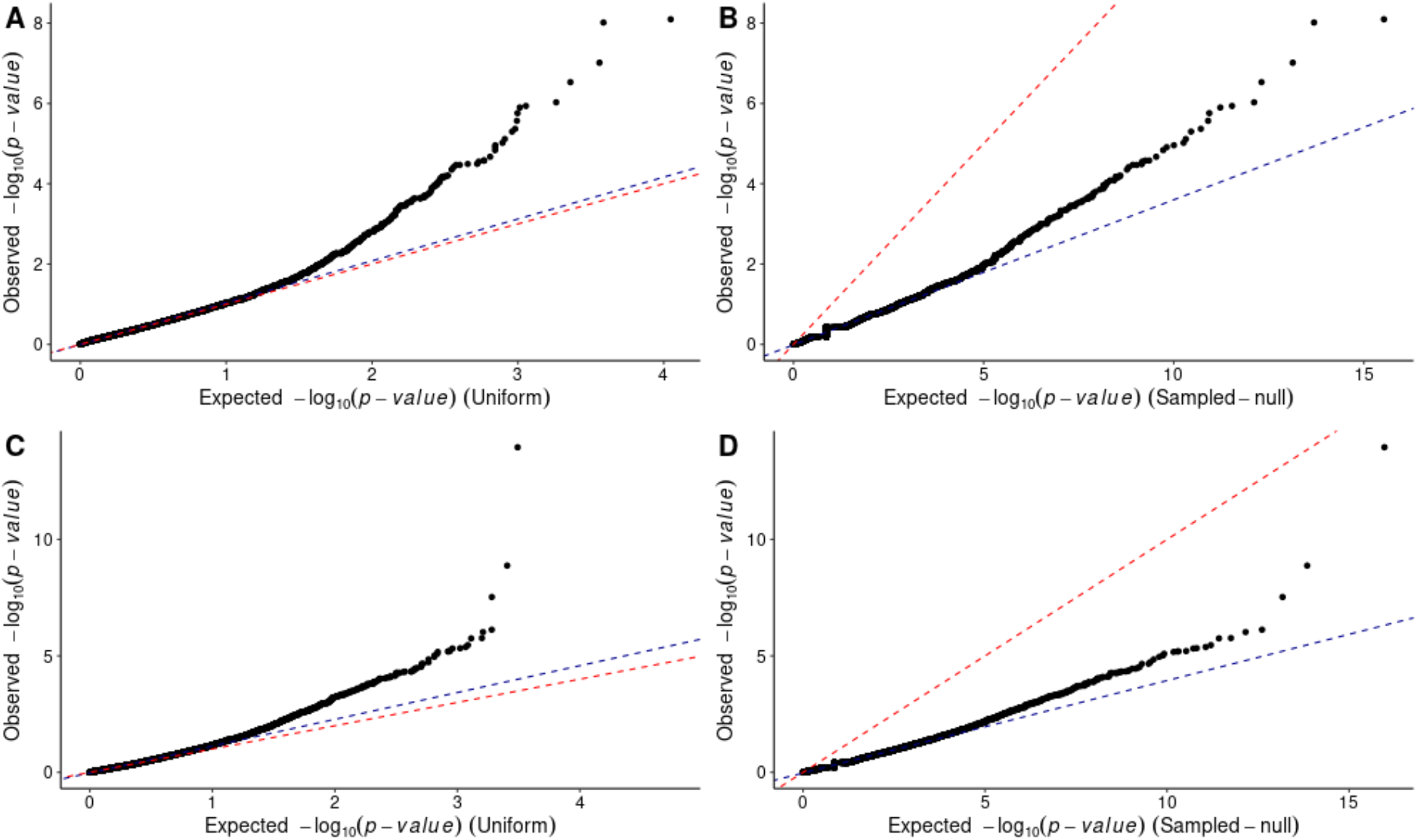
Quantile-Quantile (QQ)-plots of p-values from COBT based on qualifying variants on a cohort of 404 non-Finnish European individuals from the 1000 Genomes Project. QQ-plots represent the observed p-values (x-axis), against the expected p-values (y-axis), in minus logarithmic scale, for synonymous (top panels) and missense variants (bottom panels) with a genomic correction of p-values, using as a reference either the uniform distribution (left panels) or a sampled null distribution (right panels; **Methods**). The bisector is represented by a red dotted line. Blue dotted lines represent the linear regression of the minus log-10 scaled expected versus observed p-values based on the lower 95% quantile of points. The slope of such regressions was used to estimate the inflation factors: (A)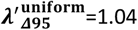; (B) 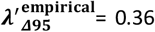; (C) 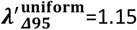; or (D) 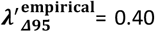. A total of 12,041 and 12,038 genes were evaluated, presenting in gnomAD at least one qualifying synonymous and missense variant, respectively. (**Methods**).

Along with the previous analyses on the cohort of 404 non-finish Europeans, we further evaluated the behaviour of two state-of-the-art methods for the identification of gene-disease associations for case-only design studies based on rare variant population frequencies: TRAPD (11) and CoCoRV (12). Both TRAPD and CoCoRV lead to p-value histograms showing a non-uniform p-value distribution both for synonymous and for missense variants (**Supplementary figure 4** and **Supplementary figure 5**), although with opposite trends. Thus, TRAPD led to an excess of significant p-values, while CoCoRV led to an excess of p-values close to 1. Furthermore, the corresponding TRAPD QQ-plots reflected a severe inflation (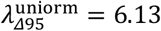 and 8.02 for synonymous and missense variants, respectively). Similarly, CoCoRV QQ-plots reflected as well important inflation levels, while less acute than in the case of TRAPD and in spite of being based on a sampled-null distribution (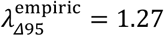 and 1.29 for synonymous and missense variants, respectively). Overall, the previous analyses on the 1000 Genomes data showed that genome-corrected COBT p-values were able to control for genomic inflation and kept type I error at low rates, as we further show in the next section. On the contrary, case-only aggregation methods such TRAPD and CoCoRV, suffered from an inflation of p-values that ultimately translated in an excess of false-positive hits, considering that no enrichments were expected for the large majority of evaluated genes in this analysis.

### Analysis of gene hits on non-Finnish European individuals from the 1000 Genomes Project

After Bonferroni multiple-testing correction setting a familywise error rate (FWER) of 5%, a total of nine (<0,01%) and eight genes (<0.08%) rejected the null hypothesis of the COBT test for synonymous and missense variants, respectively (**Table 2**). Here, rejecting the null hypothesis means that the sum of variants on a specific gene observed across the individuals from a *target* cohort (*i*.*e*., non-Finnish European from the 1000 Genomes), is statistically higher than what would be expected if the same number of individuals would be randomly selected from the *reference* cohort (*i*.*e*., non-Finnish European from gnomAD). Further inspection of the actual numbers of qualifying variants and carrier individuals for those genes reflected a clear enrichment in variants among individuals from the 1000 Genomes as compared to the expected counts inferred from gnomAD (**Table 2**). Such enrichments were not merely explained by the concentration of variants in a few carriers, as the ratio of variants per carrier ranged between 1 and 2 for most hits (**Table 2**). To better characterize the sensitivity of the test to stochastic sampling of the general population, we randomly sampled 202 out of the 404 individuals (*i*.*e*., 50%) one thousand times, and applied the COBT approach on each subset. Out of the nine and eight genes initially enriched in synonymous and missense variants, respectively, only *HRC* gene remained significant in more than 50% of the re-samplings. However, the remaining genes only rejected the null hypothesis less than 16% of the times. A partial replication is expected from the loss of statistical power associated to a lower sample size (**Supplementary figure 6**). Yet, the percentage of the *N* samplings in which hits were replicated positively correlated with the p-values observed on the total set (Wilcoxon rank sum test p-value < 2.2 × 10^6^, **Supplementary figure 7**). As a further control, the enrichments found among the 1000 random subsampling were however not explained by a severe unbalance in the retained carrier individuals between the 202 individuals considered in the corresponding COBT and the rest of the 202 individuals left aside at each sampling. Thus, mimicking a case-control setting, logistic regression analysis evaluating the association between the number of variants found in a given gene in an individual and the presence / absence of such individual among the retained split at each sampling, did not lead to any significant hit after Bonferroni correction at a FWER of 5% in more than 1% of the resampling analysis.

**Table 2:**
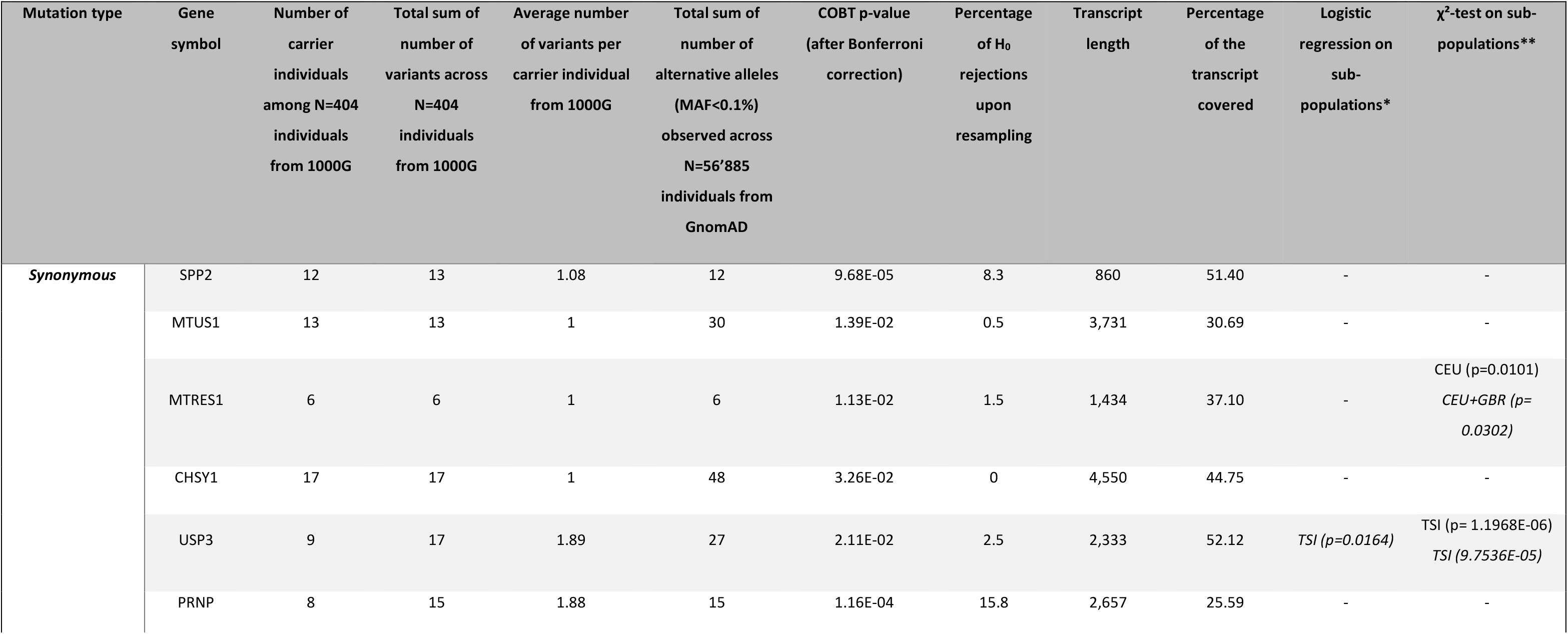
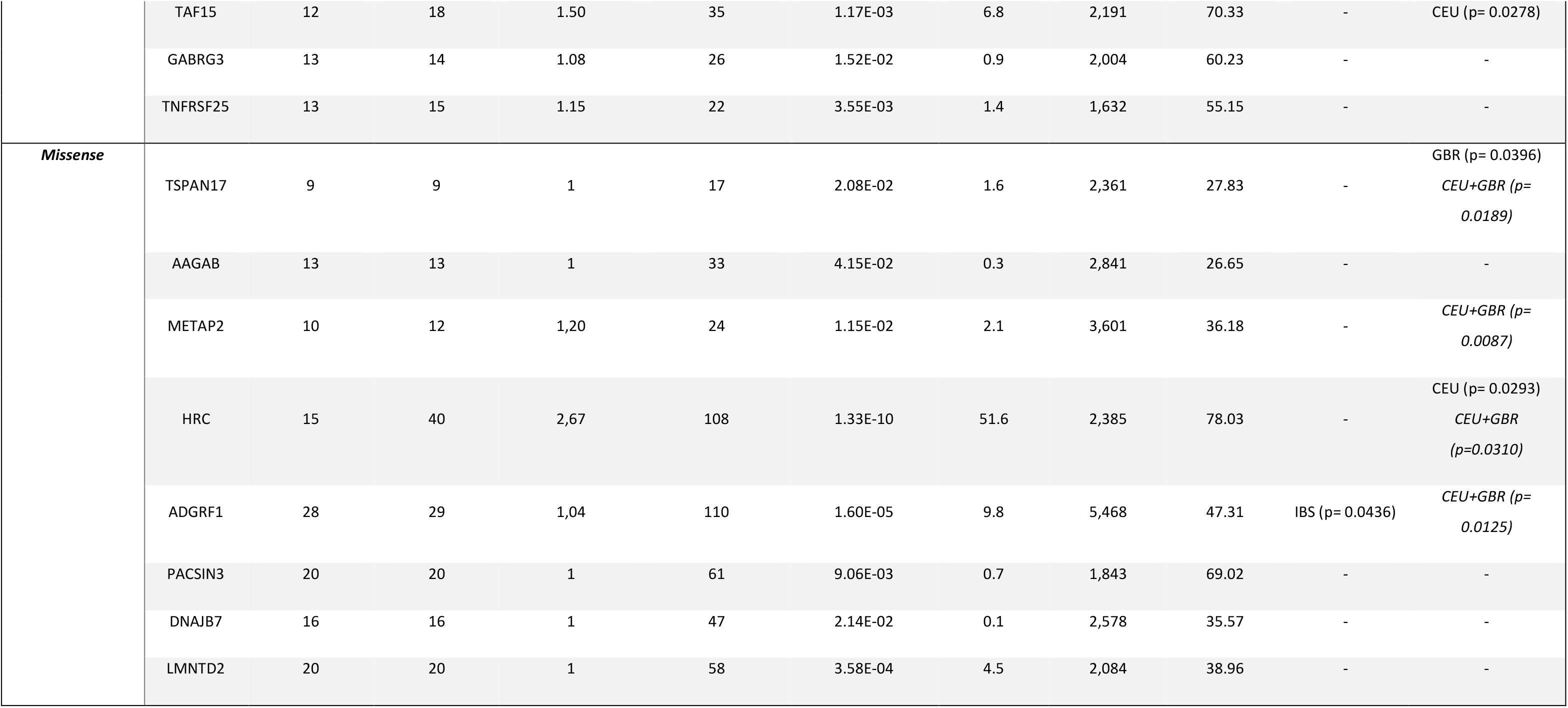
Gene hits identified by COBT on non-Finnish European individuals from the 1000 Genomes Project. The table shows the nine and eight genes rejecting the null hypothesis of COBT for synonymous and missense variants, respectively, after multiple-testing correction. The table displays the number of qualifying variants and carrier individuals found on those genes across N=404 individuals from the 1000 Genomes together with the ratio of variants per carrier. For each gene, the total sum of number of alternative alleles (MAF<0.1%) observed across N=56’885 individuals from gnomAD is shown. Random sampling of 202 out of the 404 individuals (*i*.*e*., 50%) was performed one thousand times, and the COBT approach applied on each subset. The percentage of such resampling datasets for which the gene rejected the null hypothesis is indicated in the table. In addition, the table shows the logistic regression and chi-square test p-values evaluating the association of the number of synonymous and missense variants in a specific gene with a specific subpopulation of origin of the carrier individuals, as compared to the rest of subpopulations (**Methods**). Subpopulations showing significant hits are indicated, and their Bonferroni corrected p-values (<0.05) indicated in parenthesis. Italic fonts are used when CEU and British subpopulations were jointly considered in the logistic regression and chi-square tests. CEU = Utah residents (CEPH) with Northern and Western European ancestry; TSI = Toscani in Italia; GBR = British in England and Scotland; IBS = Iberian populations in Spain.

To further characterize possible hidden factors leading to an enrichment of synonymous and missense variants in specific genes among the 1000 Genomes cohort, we inspected how carrier individuals for such variants distributed in terms of population stratification. Principal Component Analysis (PCA) based on common variants (AF>5%) of the 404 individuals actually reflected the subpopulation origin among non-Finnish Europeans, separating the Iberian and the Toscan subpopulations from the Utah residents with Northern and Western European ancestry (CEU) and the British population (**Supplementary figure 8**). The analysis of the distribution of variants across carriers showed an association with specific European sub-populations for 3 out of nine and 4 out of eight gene hits enriched in synonymous and in missense variants, respectively (Chi-squared test p-value <0.05; **Table 2**). The previous results suggest on the one hand that a fraction of the gene hits, notably those with the highest (yet significant) p-values, may originate from stochastic sampling of the general population; on the other hand, a fraction of gene hits is associated with specific European sub-populations, suggesting that such subpopulations were unevenly represented in the reference gnomAD cohort as compared to the 1000 Genomes cohort.

### Gene burden analysis of a case-only ciliopathy cohort

We then evaluated the per-gene burden of rare variants in a case-only cohort of 478 patients suffering from diverse ciliopathy types. Clinical evaluation allowed to group patients into 6 non-mutually exclusive categories according to their major phenotypes, *i*.*e*.: kidney involvement (n=381), neurological defects (n=281), skeletal defects (n=211), eye anomalies (n=182), hepatic defects (n=118) and heart defects (n=61). Yet, a wide phenotypic spectrum could be observed within each group. Ciliary exome-targeted sequencing (hereafter referred as *ciliome sequencing*) was conducted, targeting a total of 1,221 cilia-related genes (Methods). Following ACMG/AMP variant interpretation guidelines (39) clinical geneticist identified a total of 87 different causal genes (*i*.*e*., bearing homozygous or compound heterozygous variants classified as pathogenic or likely pathogenic) on n=175 patients (37% of the cohort). The rest of patients (n=303, 63%) remained genetically uncharacterized, either because pathogenic or likely pathogenic variants were found in a heterozygous state, or because no candidate variants were reported. We thus applied COBT on the global cohort of 478 patients to evaluate the per-gene burden of rare variants in order to evaluate its ability to (i) recapitulate known causal genes previously identified in the cohort, and (ii) identify novel candidate causal variants that could act as primary hits or modifiers for the genetically unsolved cases.

By applying COBT, a total of 15 genes were identified as presenting a higher burden of protein-altering variants than expected by chance (Bonferroni corrected p-value <0.05; **Table 3**; Methods). The 15 genes carrying protein-altering variants affected 194 patients, including 82 genetically resolved cases. Among those resolved cases, 21 individuals (25%) presented a causal gene previously identified through clinical assessment that is identical to the one identified by the COBT approach. Such reidentification ratio is significantly higher than what would be obtained by chance, *i*.*e*., if the same number of cilia-related gene hits (n=15) were randomly selected among the ciliome panel (Monte Carlo resampling p-value<0.003). For the remaining 75% of the patients, the causal gene that was previously identified was different from the one highlighted by COBT. In these cases, the newly identified genetic variant could play a modifying effect on the disease condition. As expected from a decrease in statistical power, the previous figures were partly reproduced when independently considering the 6 major clinical categories described above. On the contrary, no novel hits were identified in such subsets (**Table 3**). When considering missense variants only as the qualifying set, 6 genes were identified by COBT with a higher burden of variants than expected by chance (Bonferroni corrected p-value <0.05; Methods), namely CC2D1A, DNAJB13, IFT140, TTC30A, TUBA1A and WDR60. TTC30A was the only new gene hit highlighted by COBT under such setting.

**Table 3:** Summary results of the rare variant gene burden test on a case-only cohort of 478 patients suffering from diverse ciliopathy types. The table shows the 15 genes significantly enriched in protein altering variants, *i*.*e*., rejecting the null hypothesis of COBT after multiple testing correction, in a cohort of 478 patients suffering from a diverse range of ciliopathies. The table displays the gene classification as ciliopathy gene according to two reference collections (Reiter *et al*. (5) and CiliaCarta), the number of individuals carrying a qualifying protein-altering variant across N=478 patients from the in-house ciliome cohort, the number of patients with a known causal gene previously identified following ACMG/AMP variant interpretation guidelines and the number of patients p-values for which the COBT gene hit recovered the previously reported causal gene. COBT p-values are reported for each gene based on protein altering variants. COBT p-values based on missense variants are displayed in parenthesis when the gene rejected the null hypothesis considering only missense variants. Finally, COBT p-values for disease category subsets are reported when they were statistically significant for the disease categories in parenthesis.

| Gene name | Known ciliopathy gene (J.Reiter) | CiliaCarta score | Number of individuals carrying a qualifying protein-altering variant across N=478 patients from the in-house ciliome cohort | Number of patients with a known causal gene previously identified following ACMG/AMP variant interpretation guidelines | Number of patients for which the COBT gene hit recovered the previously reported causal gene | p-values | p-values for disease category subsets |
| --- | --- | --- | --- | --- | --- | --- | --- |
| **CC2D1A** | No | NA | 24 | 11 | 0 | 7.36e-3 (1.31e-2) |  |
| **CCDC40** | Yes | 3.65 | 34 | 18 | 0 | 7.95e-4 | 4.10e-4 (kidney involvement) |
| **DNAJB13** | Yes | 1.26 | 10 | 5 | 0 | 7.08e-10 (9.68e-10) | 1.45e-7 (skeletal defects)<br>1.62e-8 (neurological defects) |
| **EXOC6B** | No | -4.70 | 15 | 6 | 0 | 3.03e-2 | 4.45e-3 (kidney involvement) |
| **IFT140** | Yes | 4.86 | 45 | 26 | 8 | 1.26e-4 (1.69e-4) | 4.62e-3 (skeletal defects)<br>4.34e-3 (kidney involvement)<br>4.99e-2 (neurological defects) |
| **IFT52** | Yes | 4.86 | 14 | 10 | 0 | 5.75e-3 |  |
| **KIF3A** | Candidate | 7.27 | 15 | 8 | 0 | 1.42e-4 | 1.24e-3 (kidney involvement)<br>3.80e-3 (neurological defects) |
| **LTN1** | No | NA | 32 | 20 | 0 | 1.25e-2 | 4.53e-3 (kidney involvement) |
| **NADK2** | No | NA | 13 | 10 | 0 | 1.36e-4 | 1.53e-3 (kidney involvement) |
| **NPHP4** | Yes | 5.23 | 40 | 24 | 13 | 4.21e-2 | 1.53e-2 (kidney involvement) |
| **PDE6H** | No | NA | 8 | 4 | 0 | 6.80e-3 | 3.60e-3 (hepatic defects)<br>1.11e-3 (kidney involvement) |
| **RASAL2** | No | NA | 14 | 5 | 0 | 2.55e-3 | 5.89e-3 (skeletal defects)<br>7.77e-5 (neurological defects) |
| **SNX3** | No | NA | 7 | 4 | 0 | 8.34e-6 | 6.91e-7 (kidney involvement)<br>8.68e-4 (neurological defects) |
| **TUBA1A** | Candidate | 5.97 | 7 | 5 | 0 | 1.06e-7 (3.20e-7) | 2.79e-3 (kidney involvement)<br>9.27e-5 (neurological defects) |
| **WDR60** | Yes | 3.61 | 28 | 10 | 3 | 1.03e-3 (9.36e-4) |  |

We then evaluated the n=83 unsolved patients presenting variants in the 15 genes identified through COBT, present in heterozygous state (**Supplementary table 1**). Of them, 11 patients presented 12 variants reported as pathogenic/likely pathogenic in ClinVar (29) or disease-causing or likely disease-causing variant in HGMD (30) (**Supplementary table 1**), while an additional group of 51 patients had variants predicted as potentially pathogenic (with a CADD PHRED score higher than 15). Further manual inspection by clinical experts revealed that, among the 62 patients carrying a pathogenic or likely pathogenic variant, 22 had variants affecting genes with a functional role consistent with their observed phenotypes. Of these, 14 involved known ciliopathy genes: NPHP4 (MIM: 606966), IFT140 (MIM: 614620), or WDR60 (MIM: 615503) (**Supplementary table 2**). Notably, two patients (P5 and P14, **Supplementary table 2**) carried heterozygous variants in two different genes (*NPHP4* and *IFT140)*, either indicating a possible contribution of both variants or of one of them to the clinical phenotype. However, the segregation of the variants is unknown and should be further investigated before drawing any conclusions (40). A reanalysis of the affected individuals revealed that one of them (patient P2, **Supplementary table 1**) carry one variant in NPHP4 in association with a shared common single nucleotide variant polymorphism (rs1287637, i.e., A/T or T/T genotype, gnomAD allele frequency of 0.17) affecting splicing of NPHP4 and associated with reduced renal function (41). While this variant is currently classed as a variant of unknown significance (VUS), there is no evidence of the pathogenicity of this SNP despite the fact that it modifies the transcripts and subsequently the protein sequence (rs1287637), its combination with a pathogenic variant, if biallelic, could potentially result in a clinical NPHP4 phenotype.

An additional five patients (**Supplementary table 2**) exhibited clinical features consistent with the pathogenic or likely pathogenic variants they carried in TTC30A, a candidate gene for renal and skeletal ciliopathies (42, 43). Indeed, loss of *TTC30A* affects ciliogenesis by altering posttranslational tubulin modifications, leading to chondrodysplasia and cystic kidney disease development in xenopus models (43). Notably, patient P84 carrying a *TTC30A* variant with a missense variant with a high CADD score, presented with Saldino-Mainzer syndrome, a ciliopathy associating kidney and skeletal anomalies. Moreover, 3 patients also present with renal insufficiency associated with cerebellar vermis defects compatible with Joubert syndrome type B. Regarding the crucial role of *TTC30A* as a tubulin modulator and a component of the IFT complex, consequences of *TTC30* mutations can affect various organs including brain with features of Joubert syndrome (44). Such variants in *TTC30A* could thus constitute candidate primary hits for those patients, yet in heterozygous state. The previous results showed the capacity of COBT to recapitulate previously identified causal genes at statistically significant levels and to identify novel candidate pathogenic variants compatible with the phenotypes under investigation in unsolved cases.

## Discussion

In this work we implemented the COBT method, a statistical test for the identification of genes bearing an excess of rare variant burden in rare disease cohorts upon case-only experimental designs, *i*.*e*., when matched genomic controls are not available. To that aim, we took advantage of publicly available large exome and genome sequencing projects, allowing to obtain estimates of gene mutation rates on the general population. Comprehensive goodness-of-fit tests validated the Poisson-based statistical assumptions of the approach in terms of genetic counts dispersion, homogeneity and frequency of zeros. Furthermore, stringent filtering of genomic regions and qualifying variants together with genomic control of p-values showed COBT’s ability to keep both inflation and false positives at low rates. Yet, the COBT approach appeared as sensitive to stochastic sampling of the general population and to an uneven representation of specific subpopulations between the reference and the target cohorts, as illustrated through the analysis of a sample of non-finish Europeans from the general population. Finally, the application of COBT to the re-analysis of an inhouse cohort of ciliopathy patients genetically profiled through ciliome-sequencing showed the capacity of COBT to recapitulate previously reported causal genes in a statistically significant manner and collectively involving 21 genetically resolved patients.

In addition, COBT identified 19 unsolved patients presenting potentially-damaging variants in a heterozygous state and collectively involving 3 known ciliopathy genes (i.e., *NPHP4, IFT140, WDR60*) as well as one candidate gene for renal and skeletal ciliopathies, i.e. *TTC30A*. Expert evaluation considered that the clinical signs presented by those patients were compatible with the phenotypes and functions previously described in the literature for such genes. *TTC30A* is a component of the IFT-B complex, and like for other IFT components, variants may contribute to various manifestations among ciliopathy phenotypes (5, 45). Additionally, variants in *IFT* genes leading to syndromic ciliopathies are generally hypomorphic, allowing for compatibility with development (3, 46). Missense *TTC30A* variants identified in this study are in agreement with previously reported hypomorphic variants in *IFT-B* genes, which, while leading to syndromic ciliopathies, allow for compatibility with development (3, 45). However, for most of the patients, we were unable to identify a 2^nd^ molecular event as could be initially expected in the context of recessive inheritance, in agreement with the inheritance in most of the cases. Further investigations will be required to identify splicing defects, non-coding variants, structural rearrangements, variable number tandem repeats (VNTR), or deep intronic variants (47).

Our work is in line with recent statistical genetics tests showing the possibility of identifying gene-phenotype associations on case-only design studies by making use of aggregated population frequencies from the general population (33, 37, 48). Notwithstanding, as comprehensively reviewed (8, 10), stringent filters accounting for sequencing factors (such as depth and coverage), hidden population structure, mappability of genomic regions and the definition of qualifying variants were needed to minimize spurious signals in case-only experimental designs. Our approach represents however a conceptual novelty in the field. Thus, previously existing case-only tests such as TRAPD and CoCoRV were based on the estimation of an excess of mutated individuals for a given gene. These strategies differ from COBT where – similarly to classical gene-based burden tests for case-control designs (8) – the co-occurrence of multiple variants targeting the same gene within an individual positively contributes to the excess of actual variant burden being evaluated. Moreover, and in contrast to COBT, both TRAPD and CoCoRV presented significant inflation rates in their p-value distributions when applied to a putatively healthy cohort randomly sampled from the general population, and ultimately led to an unexpected excess of significant gene hits, which may compromise their utility for the analysis of patient cohorts.

As illustrated in a ciliopathy cohort, the COBT method can be used to identify novel gene-disease associations on retrospective studies of genetically uncharacterized patients for which no matched controls are available and as an alternative to rare variant burden tests initially developed for case-control designs. Its additive effect model assumptions may help evaluating cohorts where a large genetic heterogeneity is suspected, which could result in heterogeneous Mendelian inheritance patterns drove by dominant *de novo* variants, compound heterozygosity, oligogenic events or hypomorphic recessive variants (49). This is a meaningful assumption for ciliopathies for which, in spite of being generally considered recessive disorders -with the exception of Autosomal Dominant Polycystic Kidney Disease (ADPKD), (5) -recent reports suggest that monoallelic loss-of-function or missense variants can cause autosomal dominant retinal or renal ciliopathies (e.g., *IFT140, KIF3B, NEK8)* (40, 50– 52). Therefore, it is conceivable that some of the heterozygous variants identified in our study may contribute to the development of ciliary diseases.

Case-only burden tests were explored in this work using genes as the aggregation unit, based on their protein-coding exons. Natural extensions of the COBT approach could be proposed for the analysis of non-coding gene regions (*e*.*g*., introns, untranslated regions and promoters) either jointly or separately considered from the coding ones, as reviewed recently (53). To that aim, the rapid increase in the availability of whole genome-sequencing data from the general population may provide accurate frequency estimates of non-coding regions, as provided in the gnomAD database (13). Similarly, the COBT approach could be readily extended to the analysis of gene sets defined on the basis of a common molecular function or biological role. Thus, mutation frequencies could be aggregated to generate estimates about the number of variants that may be expected from random sampling on the general population. Gene-set based case-only burden tests could thus contribute to identify novel gene-phenotype associations that might be so far undetected due to low statistical power in small, rare disease cohorts or on diseases with a large genetic heterogeneity. In these regards, it is noteworthy that several ciliopathy patients in our study present two heterozygous compound pathogenic variants in genes encoding ciliary proteins in distinct ciliary modules, possibly suggesting synergistic effects that could contribute to phenotypic variability (54, 55). Specifically, eight patients exhibit an association of *NPHP4* variants with variants in retrograde IFT genes (*IFT140, WDR60*), which would require further investigation.

COBT may also be proposed in case-only studies based on genetically-characterized patients in the search of modifier variants associated to secondary clinical signs or severity outcomes. Such hypothesis could actually be raised in the 61 genetically resolved patients of the inhouse ciliopathy cohort evaluated in this work for which the additional variants identified by COBT could act as modifier alleles determining phenotypic variability at the individual level. This is in line with previous studies having identified genetic modifiers linked to the severity or variability of phenotypes, such as atypical neurological, retinal or kidney disease onset or progression (56–58). (59)(56)

Several limitations of the COBT approach, however, should be considered. First, COBT is limited by the fact that it does not allow to incorporate covariates such age, sex or environmental factors potentially relevant for the phenotype under investigation, in contrast to regression-based approaches for case-control analyses, which can naturally account for them (8). The association of such covariates with variant-carrier individuals of COBT gene hits may be evaluated *a posteriori* in order to disregard confounding signals. Second, COBT may be limited by low cohort sizes, which may lead to an important fraction of genes underpowered (**Supplementary figure 5**). Third, we showed that, in spite of its capacity to control for genomic inflation and keeping low false positive rates, it may still be sensitive to stochastic sampling, cryptic genetic background biases and technical artefacts. Thus, COBT gene hits should be followed up with replication analysis in independent cohorts as well as functional validations on cell lines, organoids or model organisms.

## Conclusion

The COBT method provides a statistically robust approach to identify genotype-phenotype associations through gene-based rare variant burden analysis in case-only study designs. The method is designed to account for additive effects and multiple variants within individuals, which are key features in genetically heterogeneous cohorts. COBT statistical properties were extensively validated confirming low false-positive rates and improved calibration compared to existing methods. As a validation step, the approach successfully recovered known causal genes previously identified in a cohort of ciliopathy patients and highlighted new candidate variants in genetically unresolved cases. By enabling retrospective analysis of patient cohorts that are often underused due to the absence of matched controls, COBT may provide further characterization of the architecture of human genetic diseases. COBT is implemented as an open-source software available from https://github.com/RausellLab/COBT.

## Data Availability

The COBT software developed and used in this study is openly available at https://github.com/RausellLab/COBT. Whole-genome sequencing data from the 1000 Genomes Project, used as a negative control cohort, can be accessed at http://ftp.1000genomes.ebi.ac.uk/vol1/ftp/release/20130502/. Sequencing data from the Genome Aggregation Database (gnomAD v2.1) are available at https://gnomad.broadinstitute.org/downloads. Due to patient confidentiality and ethical restrictions, individual-level sequencing data from the ciliopathy patient cohort cannot be publicly shared. Requests for access to aggregated data or additional information are subject to ethical committee approval and should be directed to the corresponding author.

https://gnomad.broadinstitute.org/downloads

http://ftp.1000genomes.ebi.ac.uk/vol1/ftp/release/20130502/

## List of abbreviations

ACMG/AMP: American College of Medical Genetics and Genomics / Association for Molecular Pathology
AF: Allele Frequency
CADD: Combined Annotation Dependent Depletion
CEU: Utah residents with Northern and Western European ancestry
ClinVar: Clinical Variant Interpretation Database
CNV: Copy Number Variant
COBT: Case-Only Burden Test
ENCODE: Encyclopedia of DNA Elements
FWER: Family-Wise Error Rate
GATK: Genome Analysis Toolkit
GBR: British in England and Scotland
HGMD: Human Gene Mutation Database
IBS: Iberian populations in Spain
IFT: Intraflagellar Transport
PCA: Principal Component Analysis
PTV: Protein Truncating Variant
QQ-plot: Quantile-Quantile Plot
SNV: Single Nucleotide Variant
SOLiD: Sequencing by Oligonucleotide Ligation and Detection
TSI: Toscani in Italia
VEP: Variant Effect Predictor
VUS: Variant of Unknown Significance

## Declarations

## Ethics approval and consent to participate

This study was conducted upon Ethical approval committee CESREES (the French ethics and scientific committee for health research), approved on September 3rd, 2020, number #2201437.

## Consent for publication

Not applicable.

## Author’s contributions

Conceived and designed the experiments: AF, SC, AR. Performed the experiments: AF. Analyzed the data: AF, SS, AR. Contributed materials/analysis tools: AF, SC, FJH, XC, NG, AB, MHH, AG, AB, YM, KB, JMR, IP, VCD, CH, MZ, TAB, SS, AR. Data curation: AB, JMR, IP, VCD, CH, TAB, SS. Wrote the paper: AF, SS, AR.

## Acknowledgments

The authors sincerely thank the affected individuals and their families for participation, and the clinicians, for their contribution to this study. We are grateful to Sandra Colas (URC Necker) and Johanna Nunez Ordonez for their contribution. We greatly acknowledge members of the bioinformatic and genomic facilities of the Imagine Institute.

