## Supplementary Figures for "COBT: A gene-based rare variant burden test for case-only study designs using aggregated genotypes from public reference cohorts"

### Supplementary tables:

The two supplementary tables can be found in the attached file : "CaseOnly_Manuscript_20250520_Suppl_tables_AF.xlsx".

**Supplementary table 1. Manual assessment of phenotypic compatibility for gene variants identified by COBT in n = 83 unsolved ciliopathy patients.** The table presents the expert manual evaluation of the phenotypic compatibility between each patient's clinical presentation (i.e., observed symptoms) and the gene hits identified through COBT analysis. It includes the clinical significance of the detected variants, based on annotations from ClinVar, HGMD, and CADD scores. White and grey alternating rows are used to group variants belonging to the same patient. Where available, additional information supporting compatibility—such as ORPHA codes or the presence of additional variants—is included. The "Disease reported" column summarizes literature-based evidence regarding the pathogenicity of the identified variants. Patient-gene pairs labelled as "compatible" indicate a match between the patient's phenotype and the known clinical manifestations associated with the gene according to the literature. Pairs marked as "possibly compatible" refer to cases where the phenotype falls within a gene's known broader clinical spectrum or within "compatible" genes but associated with variants of low-confidence significance (e.g., benign or uncertain ClinVar annotations and low CADD scores). Of note, while PDE6H was identified as a COBT hit, no variants with associated CADD scores were found for this gene; therefore, it was not subjected to manual validation.

**Supplementary table 2. Known and candidate ciliopathy gene hits identified by COBT with pathogenic or probably pathogenic variants and their phenotypic compatibility in ciliopathy patient carriers.** The table shows the phenotypic compatibility assessed by clinical experts between the patient's phenotype (*i.e.*, clinical symptoms) and specific gene hits identified by COBT (*WDR60*, *NPHP4*, *IFT140* and *TTC30A*) with pathogenic variants (assessed by HGMD, ClinVar or with CADD PHRED score > 15, **Methods**). TTC30A was detected by COBT using only missense variants in the qualifying set. White and grey rows highlight variants of the same genes. Patients who contributed variants in multiple COBT hit genes are indicated with a symbol between parentheses. Additional information to the compatibility is provided when available (*i.e.*, ORPHA code, additional variant etc). The "disease reported" column provides information from the literature about the pathogenicity of the variants.

### Supplementary figures:

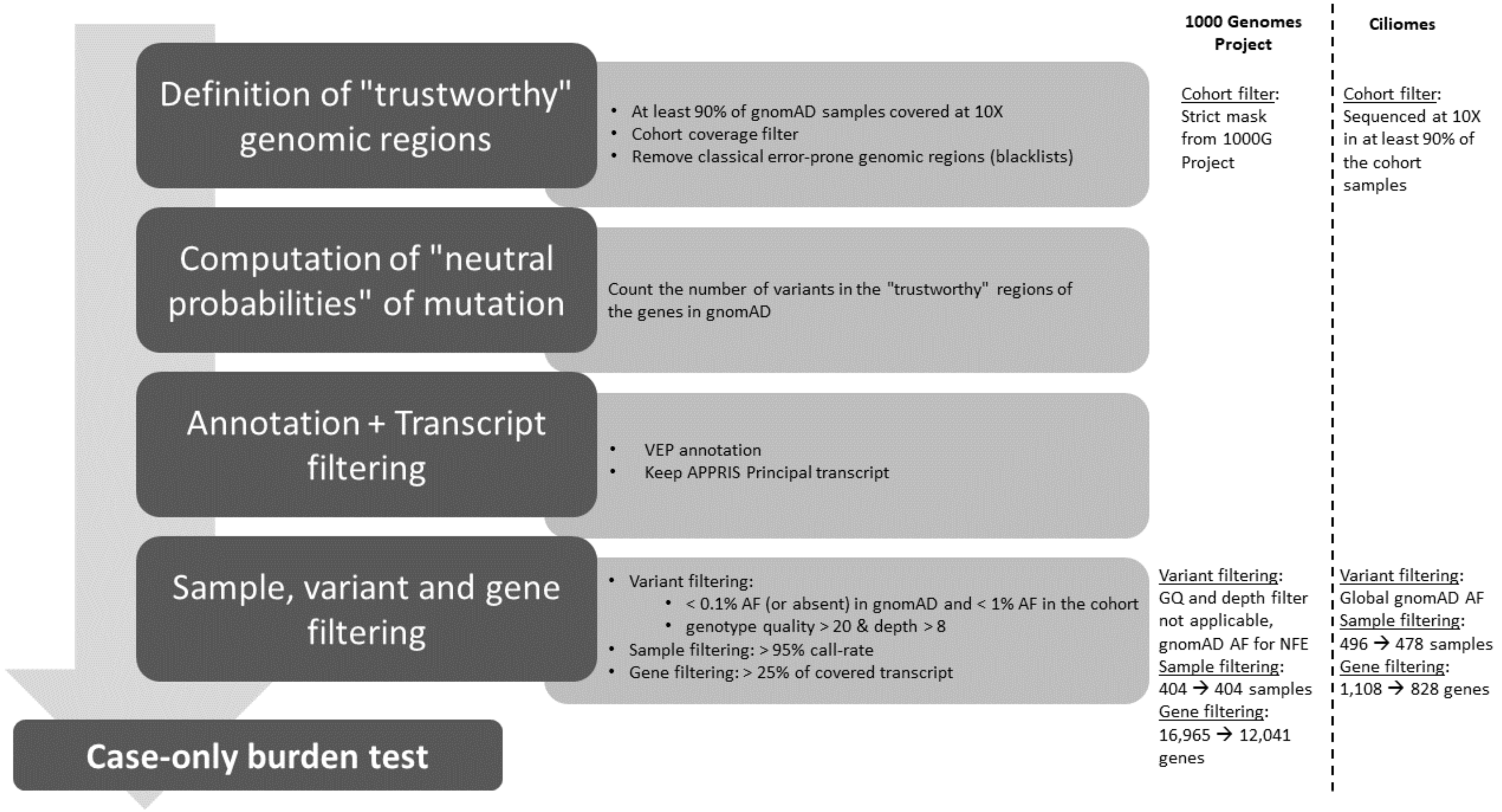


**Supplementary figure 1:** Schematic representation of the genomic and sample filtering as well as genetic variant annotation and filtering performed on the two target cohorts considered throughout the study: whole-genome sequencing data from non-Finnish European individuals from the 1000 Genomes project (15) and ciliary exome-targeted sequencing (**Methods**).

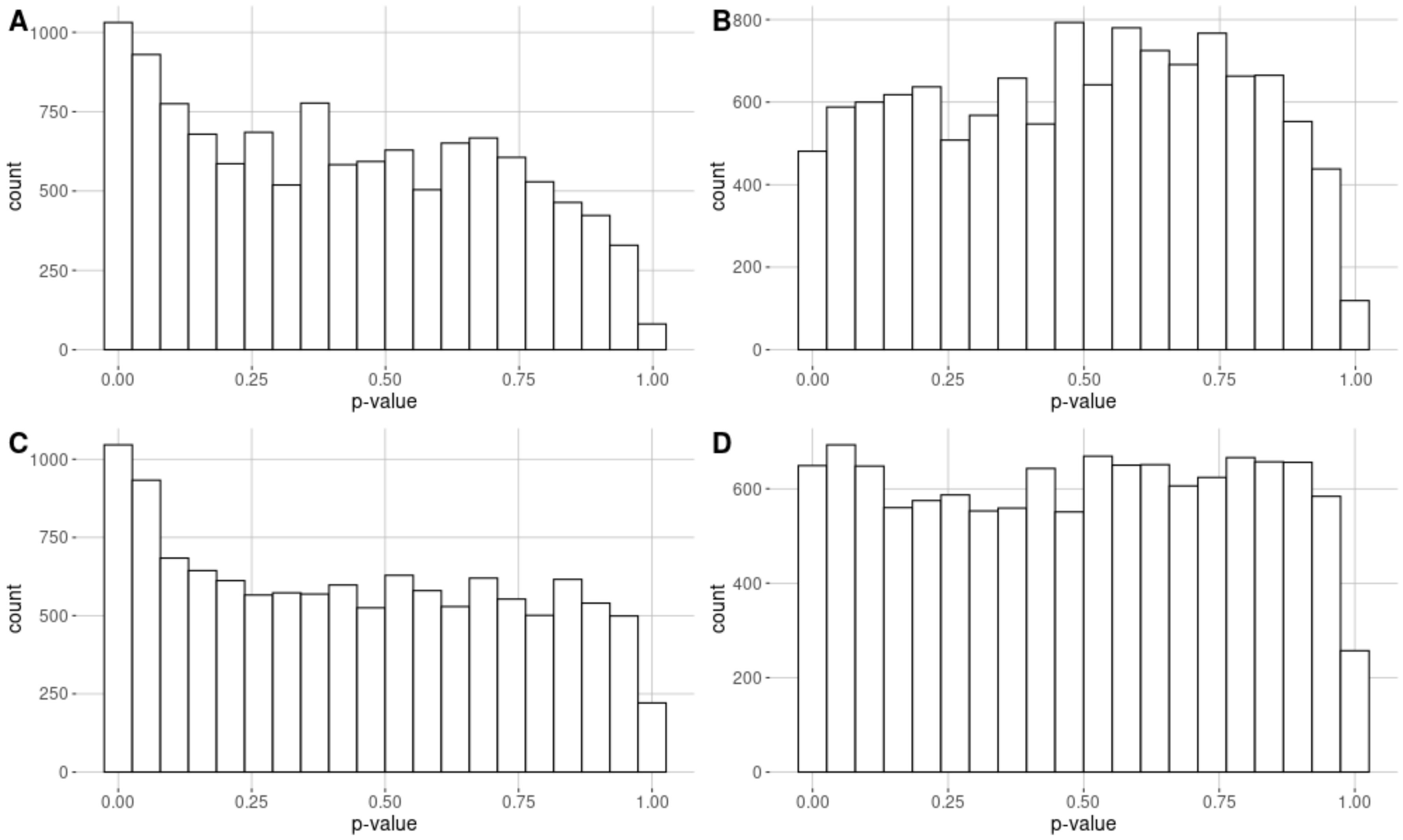


**Supplementary figure 2: Distribution of p-values from COBT on a cohort of 404 non-Finnish European individuals from the 1000 Genomes Project.** Histograms show the p-value distribution both for synonymous (**top** panels) and for missense (**bottom** panels) variants, without (**left** panels) or with a genomic correction of p-values (**right** panels). A total of 12,041 and 12,038 genes were evaluated, presenting in gnomAD at least one qualifying synonymous and missense variant, respectively (**Methods**).

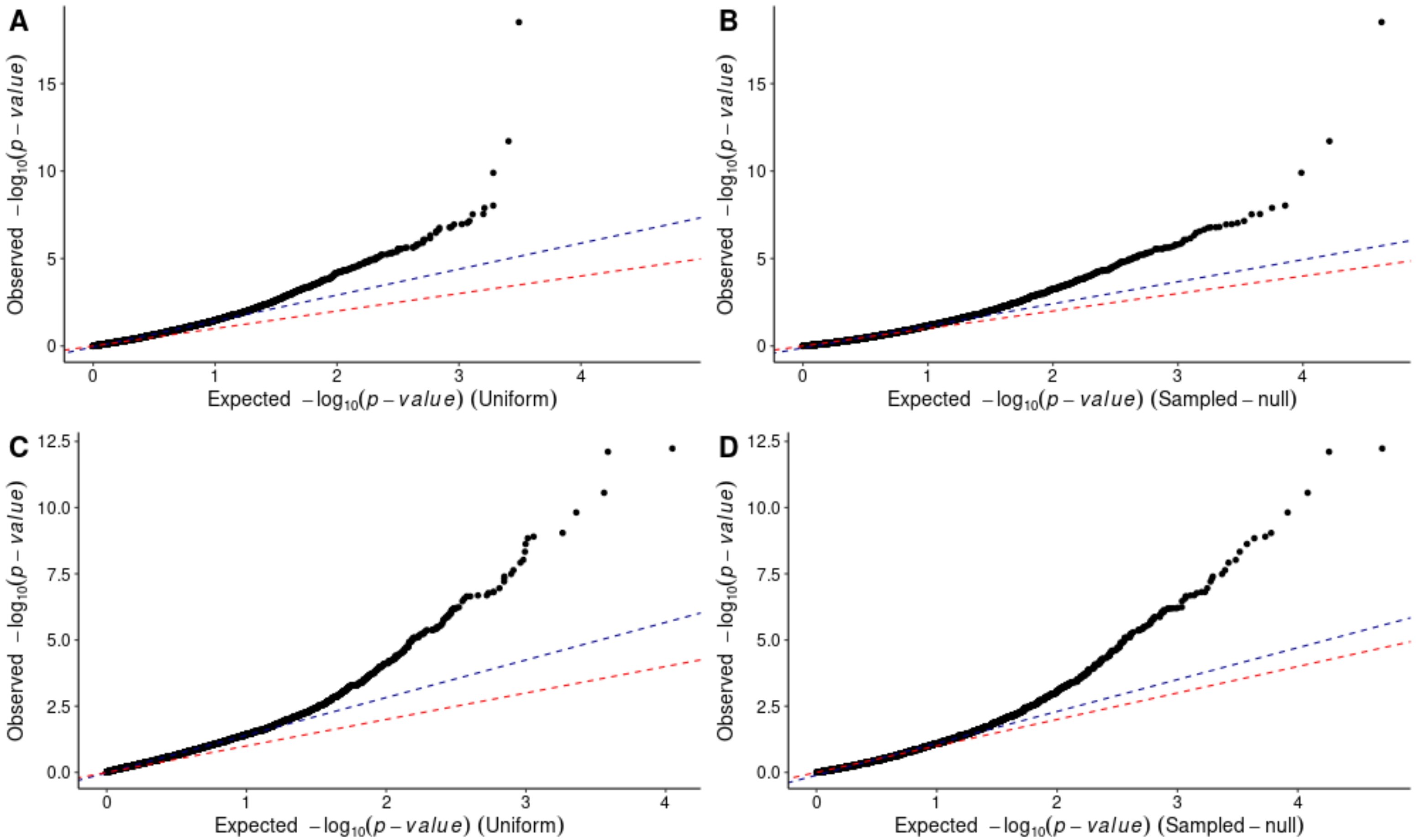


**Supplementary figure 3: Quantile-Quantile (QQ)-plots of p-values from COBT based on qualifying variants on a cohort of 404 non-Finnish European individuals from the 1000 Genomes Project.** QQ-plots represent the observed p-values (x-axis), against the expected p-values (y-axis), in minus logarithmic scale, for synonymous (**top** panels) and missense variants (**bottom** panels) without a genomic correction of p-values, using as a reference either the uniform distribution (**left** panels) or a sampled null distribution (**right** panels; Methods). The bisector is represented by a red dotted line. Blue dotted lines represent the linear regression of the minus log-10 scaled expected versus observed p-values based on the lower 95% quantile of points. The slope of such regressions was used to estimate the inflation factors: (**A**) $\lambda_{\Delta 95}^{\mathbf{uniform}}$ =1.42; (**B**) $\lambda_{\Delta 95}^{\mathbf{empirical}}$= 1.20; (**C**) $\lambda_{\Delta 95}^{\mathbf{uniform}}$ =1.48; (**D**) $\lambda_{\Delta 95}^{\mathbf{empirical}}$= 1.26. A total of 12,041 and 12,038 genes were evaluated, presenting in gnomAD at least one qualifying synonymous and missense variant, respectively (**Methods**).

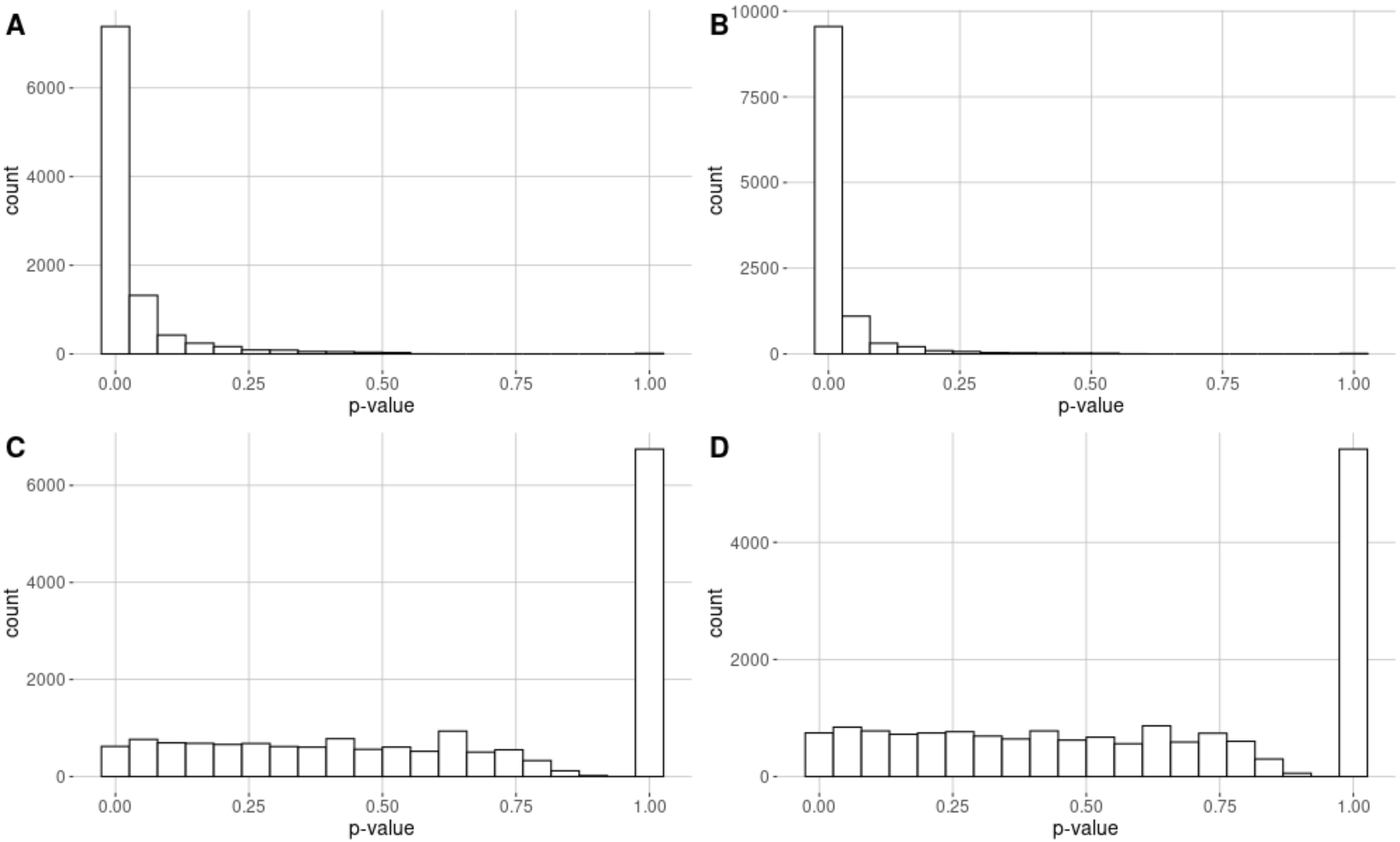


**Supplementary figure 4: Distribution of p-values from the TRAPD and CoCoRV tests on a cohort of 404 non-Finnish European individuals from the 1000 Genomes Project.** Histograms show the raw p-value distributions (*i.e.*, uncorrected p-values) both for synonymous (**top** panels) and for missense (**bottom** panels) variants, from the TRAPD (**left** panels) and CoCoRV tests (**right** panels). A total of 9,900 and 11,496 genes carrying at least one synonymous and missense variant respectively were evaluated in TRAPD test and 18,283 genes carrying at least one qualifying variant for both synonymous and missense variants were evaluated in CoCoRV test (**Methods**).

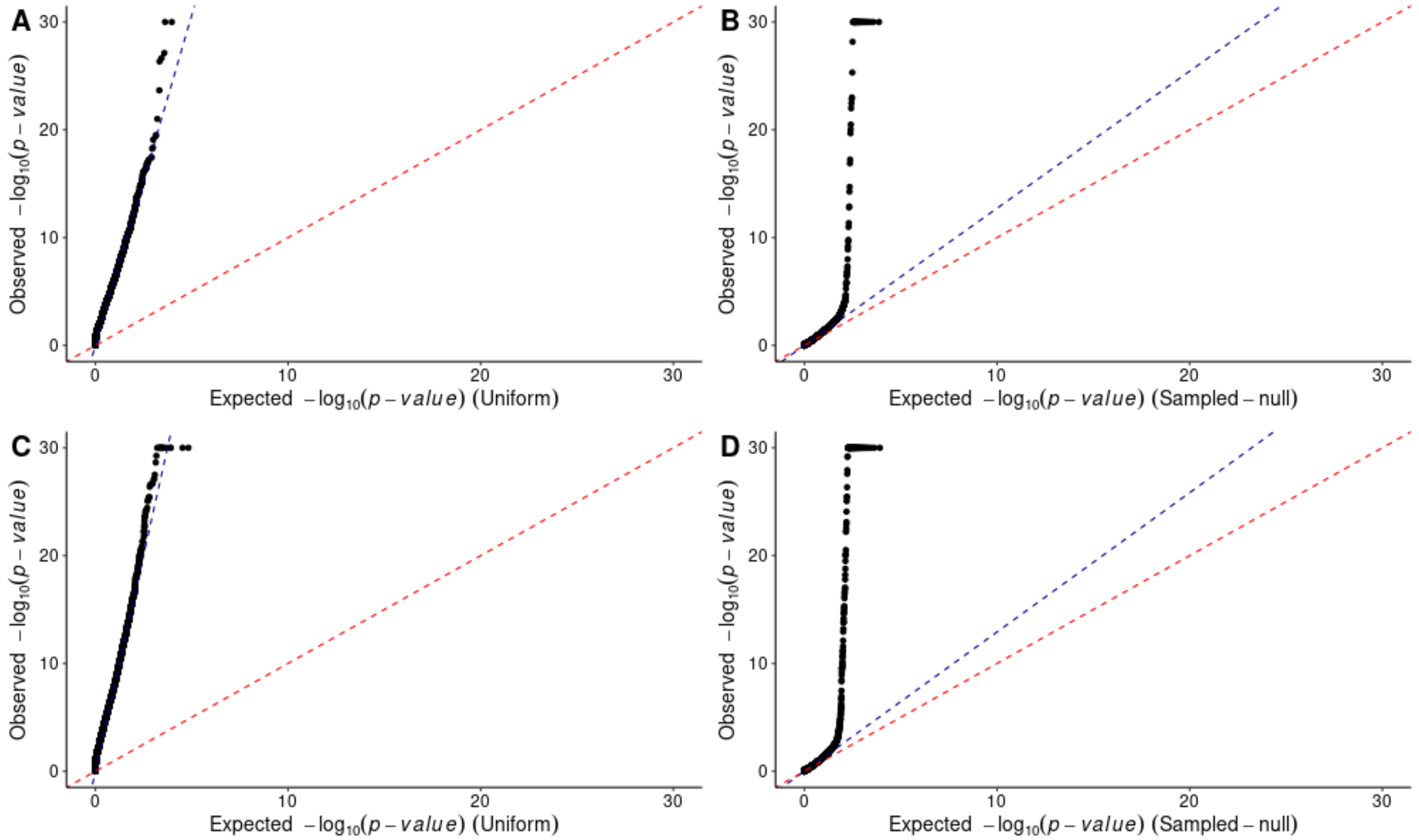


**Supplementary figure 5: Quantile-Quantile (QQ)-plots of p-values from the TRAPD and CoCoRV tests based on qualifying variants on a cohort of 404 non-Finnish European individuals from the 1000 Genomes Project.** QQ-plots represent the observed raw p-values (x-axis), against the expected p-values (y-axis), in minus logarithmic scale, from the TRAPD (**left** panels) and CoCoRV tests (**right** panels), based on synonymous (**top** panels) and missense variants (**bottom** panels). Following the original implementations of both tests, the uniform distribution was used as a reference null distribution in the case of TRAPD, while a sampled null distribution was used in the case of CoCoRV (**right** panels; **Methods**). The bisector is represented by a red dotted line. Blue dotted lines represent the linear regression of the minus log-10 scaled expected versus observed p-values based on the lower 95% quantile of points. For the sake of visualization, minus log-10 p-values higher than 30 were capped to a value of 30. The slope of such regressions was used to estimate the inflation factors: (**A**) $\boldsymbol{\lambda}_{\boldsymbol{\Delta 95}}^{\mathbf{TRAPD}}$ = 6.13; (**B**) $\boldsymbol{\lambda}_{\boldsymbol{\Delta 95}}^{\mathbf{CoCoRV}}$= 1.27; (**C**) $\boldsymbol{\lambda}_{\boldsymbol{\Delta 95}}^{\boldsymbol{TRAPD}}$=8.02; or (**D**) $\boldsymbol{\lambda}_{\boldsymbol{\Delta 95}}^{\mathbf{CoCoRV}}$= 1.29. A total of 9,900 and 11,496 genes carrying at least one synonymous and missense variant respectively were evaluated in TRAPD test and 18’283 genes carrying at least one qualifying variant for both synonymous and missense variants were evaluated in CoCoRV test (**Methods**).

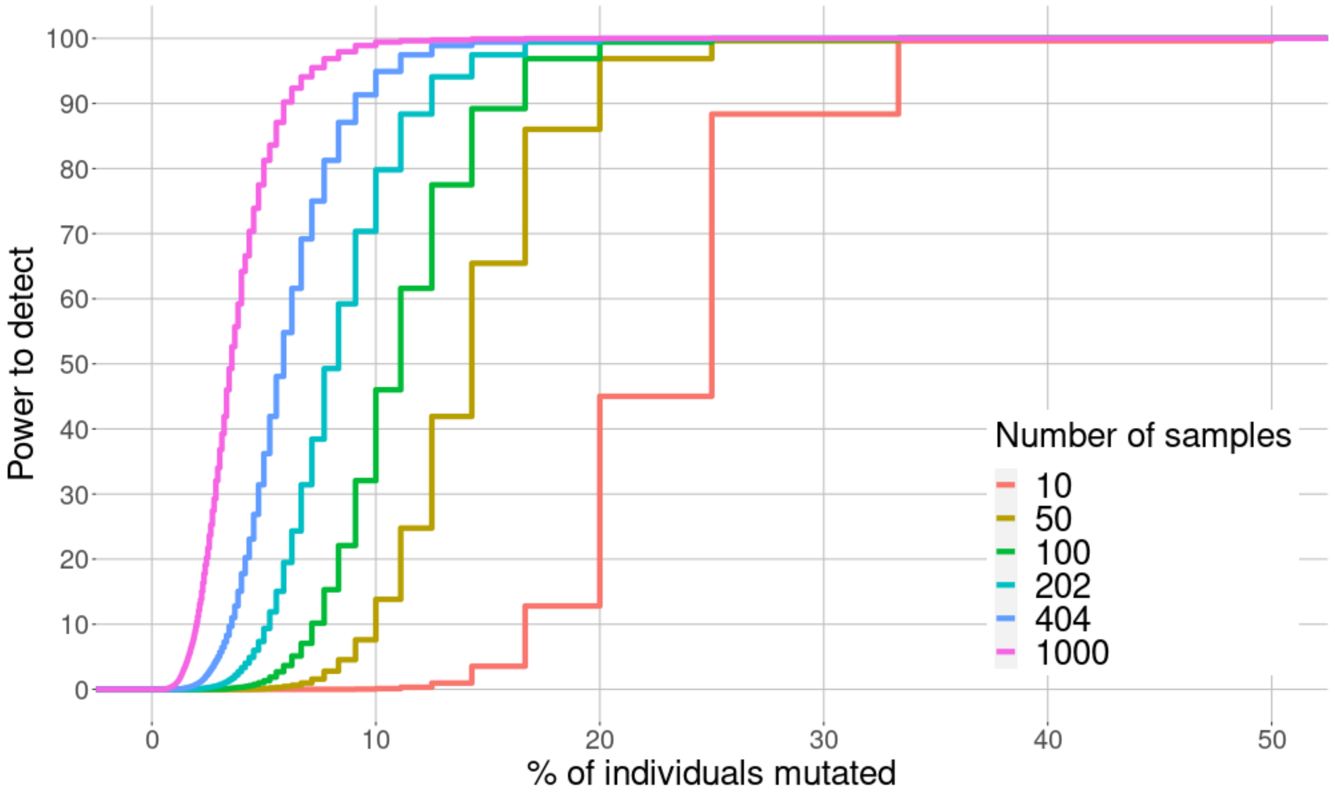


**Supplementary figure 6: Power analysis of COBT.** Impact of the sample size on the power of the test to detect genes carrying a burden of variants. The plot represents the percentage of mutated individuals in the cohort (x-axis) against the power to detect genes carrying a rare variant burden (y-axis) for different cohort sizes represented by each curve.

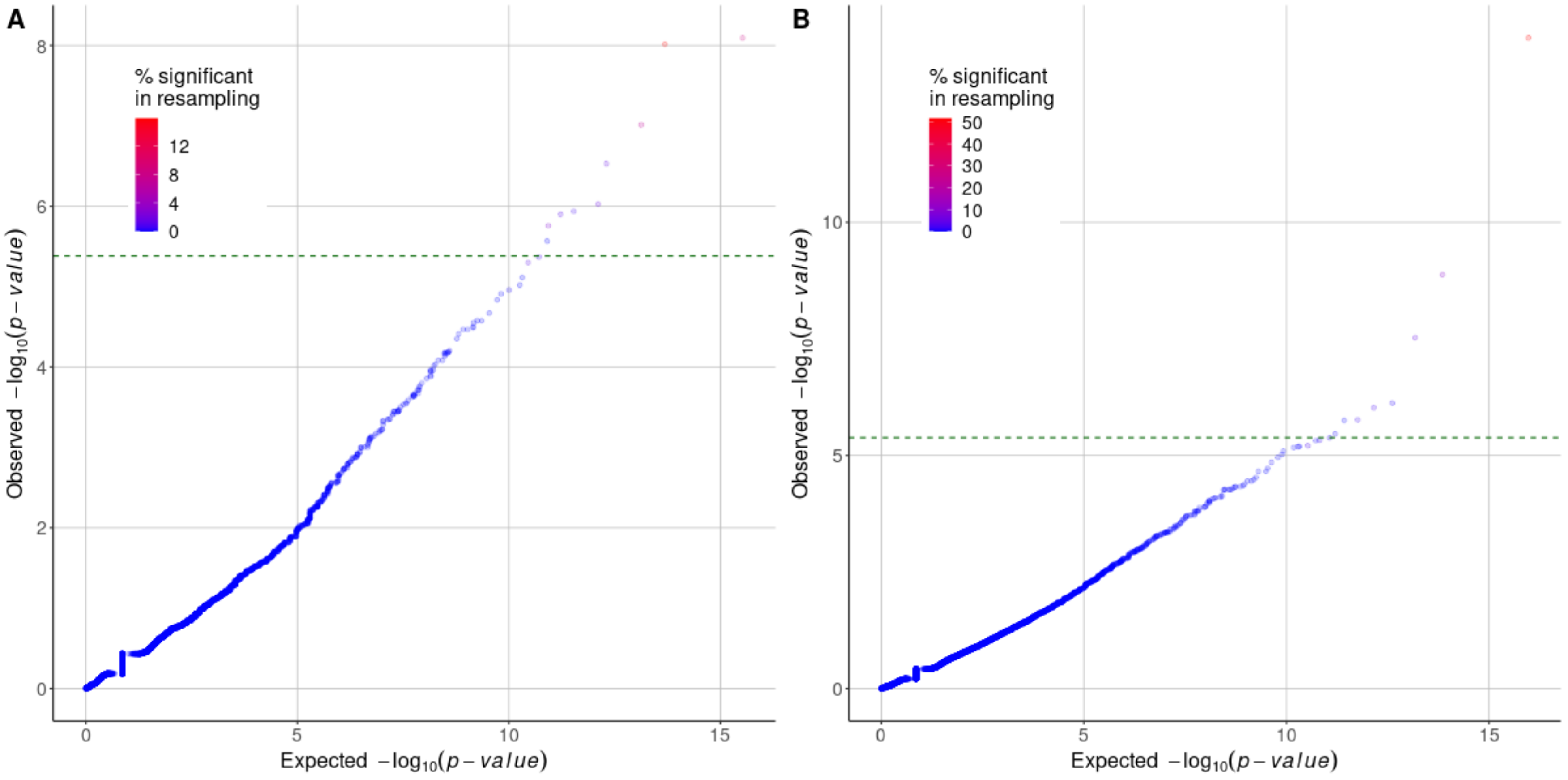


**Supplementary figure 7: Characterization of the sensitivity of the COBT test to stochastic sampling of the general population.** The figure shows the Quantile-Quantile (QQ)-plots of p-values from COBT based on qualifying synonymous (**A**) and missense (**B**) variants on a cohort of 404 non-Finnish European individuals from the 1000 Genomes Project. QQ-plots represent the observed p-values (x-axis), against the expected p-values (y-axis), in minus logarithmic scale, for synonymous variants, with a genomic correction of p-values using as a reference a sampled null distribution (**Methods**). Random sampling of 202 out of the 404 individuals (*i.e.*, 50%) was performed one thousand times for each gene, and the COBT approach was applied on each subset. The percentage of such resampling dataset for which the gene rejected the null hypothesis is indicated in a color scale ranging from blue (0%) to red (14 and 50% for synonymous and missense variants, respectively). The percentage of the N samplings in which hits were replicated positively correlated with the p-values observed on the total set (Wilcoxon rank sum test p-value < $2.2 \times 10^{6}$). The green dotted line represents the raw p-value significance threshold leading to Bonferroni corrected p-values < 0.05. A total of 12,041 and 12,038 genes were evaluated, presenting in gnomAD at least one qualifying synonymous and missense variant, respectively (**Methods**).

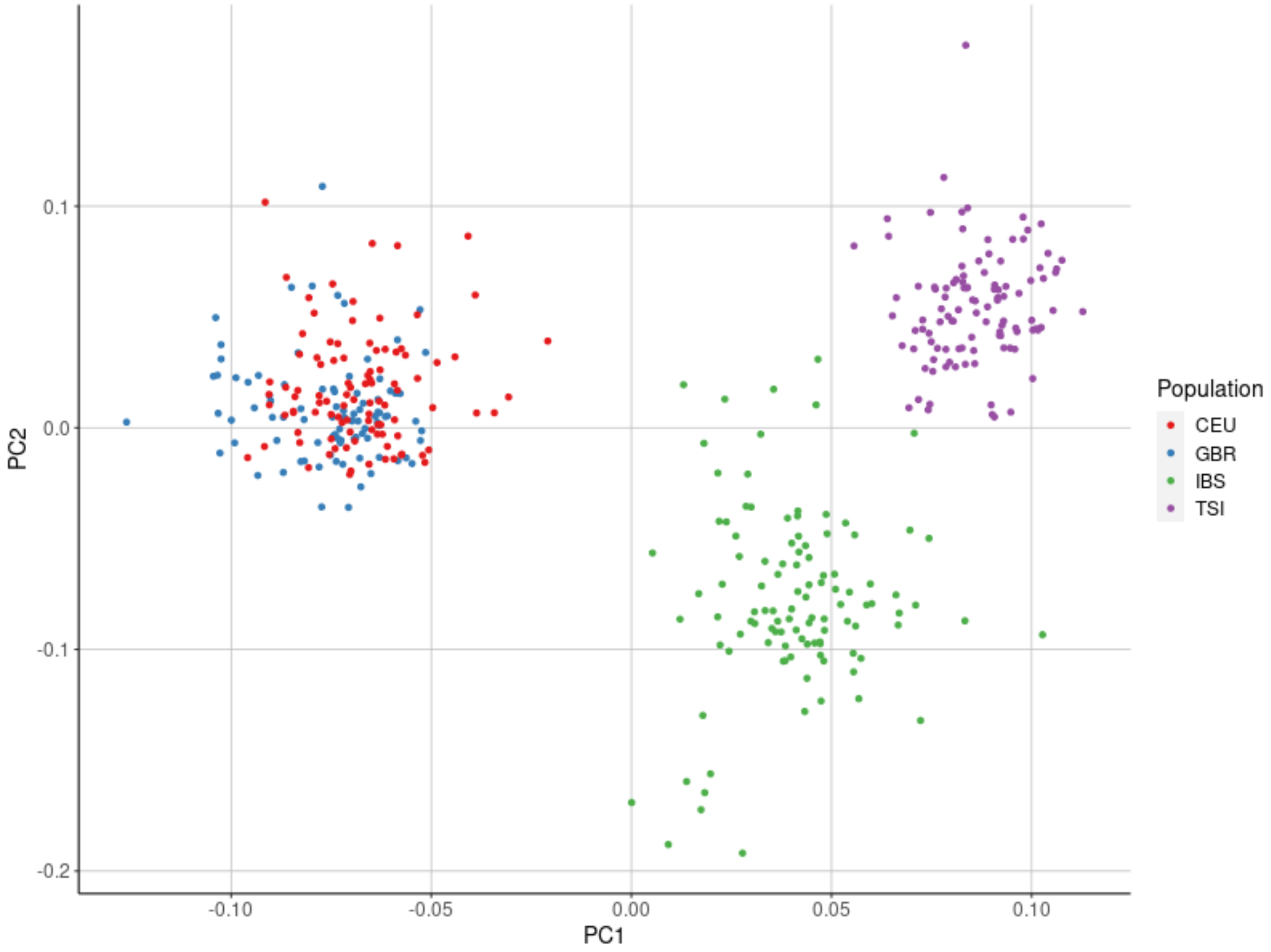


**Supplementary figure 8: Principal Component Analysis of non-Finnish European individuals from the 1000 Genomes Project.** The figure represents the projection on the first two principal components of 404 non-Finnish European individuals based on n= 9,735,687 common genetic variants (minor allele frequency > 0.01, call-rate ≥ 0.95 and HWE p-value ≥ $1.10^{-6}$) projected on the first two principal components, coloured by subpopulation of origin. CEU = Utah residents (CEPH) with Northern and Western European ancestry; TSI = Toscani in Italia; GBR = British in England and Scotland; IBS = Iberian populations in Spain.
